# Validation and culture adaptation of the Arabic version of the Rapid Eating Assessment for Participants - Shortened Version (REAP-S): A Confirmatory Factor Analysis and Rasch Analysis Study

**DOI:** 10.64898/2026.10.15.26365598

**Authors:** Talal M. Albarrak, Walid Al-Qerem, Seirsa N. AL-Quraan, Abdulhafith Y. Alharbi, Osama Alkouri

**Author notes:** Corresponding Author: Seirsa Nasser AL-Quraan, PhD, RN, Clinical Nursing Department- Faculty of Nursing- Jerash Private University, Jerash, 26150 Jordan.

## Abstract

**Aim:** To translate, culturally adapt, and psychometrically validate the Arabic version of the REAP-S among Arabic speaking young adults using Confirmatory Factor Analysis (CFA) and Rasch Measurement Theory (RMT).

**Design:** A cross-sectional methodological psychometric validation study.

**Methods:** The study included 500 undergraduate and postgraduate nursing students recruited from Saudi universities during the 2025–2026 academic year. The REAP-S was translated into Arabic using standardized forward backward translation procedures and cross-cultural adaptation guidelines. Participants completed the Arabic REAP-S through an online self-administered questionnaire. Construct validity was evaluated using CFA with robust estimators, assessing the Comparative Fit Index (CFI), Tucker-Lewis Index (TLI), Root Mean Square Error of Approximation (RMSEA), and Standardized Root Mean Square Residual (SRMR). Rasch Measurement Theory was applied to examine item fit, response category functioning, person item targeting, and person reliability. We assessed internal consistency using Cronbach’s alpha and the Rasch Person Separation Index (PSI).

**Results:** Psychometric analyses included 490 participants with a median age of 20 years (IQR = 19–22), of whom 54.1% were female. CFA supported the hypothesized one-factor structure and showed good model fit (CFI = 0.984, TLI = 0.978, RMSEA = 0.076, SRMR = 0.061). Standardized factor loadings ranged from 0.429 to 0.768, and internal consistency was good (Cronbach’s α = 0.863). Rasch analysis confirmed the scale’s unidimensionality, with satisfactory person reliability (Expected A Posteriori [EAP] = 0.872; Weighted Likelihood Estimate [WLE] = 0.848). All items demonstrated acceptable infit and outfit mean square statistics (0.826–1.311 and 0.866–1.283, respectively), with ordered response thresholds and adequate person-item targeting. Overall, the Arabic REAP-S effectively discriminated between different levels of dietary behavior among Arabic-speaking young adults.

**Conclusion:** The Arabic REAP-S demonstrated strong validity and reliability, supporting its unidimensional structure and appropriate psychometric performance. Evidence from both CFA and Rasch analysis confirmed acceptable model fit, good internal consistency, and adequate item functioning. The instrument provides a brief and practical tool for assessing dietary behaviors among Arabic-speaking young adults in clinical and research settings.

**Implications for the Profession and/or Patient Care:** The Arabic REAP-S provides nurses and other healthcare professionals with a brief, standardized tool for rapidly assessing dietary behaviors and identifying individuals at nutritional risk. Its integration into routine clinical and community practice may support early dietary screening, guide nutrition education, and strengthen evidence based preventive care for diet related chronic diseases.

## Introduction

Dietary behaviors are key determinants of health and are strongly associated with the prevention and management of non-communicable diseases (NCDs), including obesity, cardiovascular diseases, and type 2 diabetes (Collaborators, 2020; Afshin et al., 2023). The increasing burden of diet-related diseases has become a major public health concern worldwide, emphasizing the need for effective strategies to assess and improve dietary habits (Afshin et al., 2023). Among young adults, dietary behaviors are often characterized by unhealthy eating patterns, meal skipping, and increased consumption of energy dense foods, which may contribute to adverse health outcomes later in life (Deliens et al., 2014; Pendergast et al., 2016). Accurate assessment of dietary behaviors is essential for identifying individuals at nutritional risk, evaluating dietary interventions, and informing public health policies. Although comprehensive dietary assessment methods can provide detailed information about food intake, they are often time-consuming and impractical for routine clinical and community use. Consequently, brief dietary screening tools have become increasingly important due to their ease of administration and ability to rapidly identify unhealthy dietary patterns (Segal-Isaacson et al., 2004; Shankar et al., 2023).

The Rapid Eating Assessment for Participants–Shortened Version (REAP-S) is a brief self-administered dietary screening questionnaire developed to assess eating behaviors associated with dietary quality and healthy eating practices (Segal-Isaacson et al., 2004). The instrument has demonstrated acceptable validity and reliability and has been used in various healthcare and research settings to support nutritional assessment and counseling (Segal-Isaacson et al., 2004). More recently, an updated version of the REAP-S has shown strong psychometric properties, further supporting its usefulness as a rapid dietary screening tool (Shankar et al., 2023). Despite the growing need for practical dietary assessment instruments in Arabic speaking populations, few brief dietary screening tools have undergone rigorous translation, cultural adaptation, and psychometric evaluation. Cross-cultural adaptation is a critical step in ensuring that translated instruments maintain conceptual equivalence and accurately reflect the intended constructs within different linguistic and cultural contexts (Boateng et al., 2018; Lin et al., 2025). Establishing the validity and reliability of culturally adapted instruments is necessary before their implementation in research and clinical practice (Boateng et al., 2024).

Contemporary recommendations for instrument validation advocate the use of both classical and modern psychometric approaches to provide comprehensive evidence regarding measurement quality (Furr, 2021; Mokkink et al., 2023). Confirmatory factor analysis (CFA) is widely used to evaluate construct validity and determine whether observed data support a hypothesized factor structure. Model fit is commonly assessed using indices such as the Comparative Fit Index (CFI), Tucker–Lewis Index (TLI), Root Mean Square Error of Approximation (RMSEA), and Standardized Root Mean Square Residual (SRMR) (Kenny et al., 2015). In addition, Rasch Measurement Theory provides valuable information regarding item functioning, response category performance, measurement precision, and person–item targeting, offering insights that complement traditional psychometric analyses (Bond & Fox, 2013; Tennant & Conaghan, 2007; Boone et al., 2021; Petrillo et al., 2024).

Therefore, the present study aimed to translate and culturally adapt the Rapid Eating Assessment for Participants–Shortened Version (REAP-S) into Arabic and to evaluate its psychometric properties among Arabic-speaking young adults. Specifically, the study examined the internal consistency, construct validity, and measurement properties of the Arabic REAP-S using Confirmatory Factor Analysis and Rasch Measurement Theory. Establishing a valid and reliable Arabic version of the REAP-S may provide researchers and healthcare professionals with a practical instrument for assessing dietary behaviors and supporting nutrition-related interventions within Arabic-speaking populations.

### Study aims

The study aimed to translate and culturally adapt the Rapid Eating Assessment for Participants–Shortened Version (REAP-S) into Arabic and to assess its psychometric performance among Arabic speaking young adults. It evaluated construct validity, internal consistency, item level functioning, and overall measurement properties using Confirmatory Factor Analysis (CFA) and Rasch Measurement Theory.

### Theoretical Framework

The present study is based on the theoretical premise that dietary behavior is a measurable health related construct that influences the prevention and management of non-communicable diseases (NCDs). Dietary patterns, food choices, and eating habits are recognized as major determinants of health outcomes, particularly obesity, cardiovascular diseases, and type 2 diabetes (Afshin et al., 2023; Collaborators, 2020). Consequently, accurate assessment of dietary behavior is essential for identifying nutritional risk, monitoring dietary practices, and informing health promotion interventions (Deliens et al., 2014; Pendergast et al., 2016).

The Rapid Eating Assessment for Participants–Shortened Version (REAP-S) was developed as a brief dietary screening instrument to assess eating behaviors associated with dietary quality and adherence to healthy eating recommendations (Segal-Isaacson et al., 2004). The instrument is grounded in the assumption that responses to individual questionnaire items reflect an underlying latent construct of healthy dietary behavior. In psychometric theory, latent constructs are not directly observable but can be inferred through individuals’ responses to a set of related indicators (Furr, 2021). Therefore, the validity of the REAP-S depends on the extent to which its items accurately represent and measure the construct of dietary behavior.

The current study is also informed by the principles of cross-cultural adaptation and measurement equivalence. According to established guidelines, the adaptation of a health measurement instrument involves more than linguistic translation; it requires the preservation of conceptual, semantic, and cultural equivalence between the original and translated versions (Boateng et al., 2018; Boateng et al., 2024). Cross-cultural adaptation seeks to ensure that respondents from different cultural and linguistic backgrounds interpret questionnaire items in a manner consistent with the original instrument’s intended meaning (Lin et al., 2025). Without adequate adaptation and validation, measurement error may arise, potentially compromising the accuracy and interpretability of the instrument (Boateng et al., 2018).

To evaluate the psychometric performance of the Arabic REAP-S, the study draws upon both Classical Test Theory (CTT) and Rasch Measurement Theory (RMT), which are increasingly recommended as complementary approaches in instrument validation research (Furr, 2021; Mokkink et al., 2023). Within the CTT framework, construct validity is examined through Confirmatory Factor Analysis (CFA). CFA assesses whether the observed relationships among questionnaire items are consistent with a hypothesized latent factor structure and provides evidence regarding the extent to which the instrument measures the intended construct (Kenny et al., 2015). Model adequacy is commonly evaluated using fit indices such as the Comparative Fit Index (CFI), Tucker–Lewis Index (TLI), Root Mean Square Error of Approximation (RMSEA), and Standardized Root Mean Square Residual (SRMR) (Kenny et al., 2015).

In addition to construct validity, reliability is a fundamental component of measurement quality. Internal consistency reliability reflects the degree to which questionnaire items measure the same underlying construct and produce consistent scores across respondents (Furr, 2021). Adequate internal consistency provides evidence that the items function collectively as indicators of dietary behavior and supports the interpretation of total scale scores.

Complementing the CTT framework, Rasch Measurement Theory offers a modern psychometric approach that evaluates the interaction between person ability and item difficulty on a common measurement continuum (Bond & Fox, 2013; Boone et al., 2021). Rasch analysis examines whether individual items conform to model expectations, whether response categories function as intended, and whether the instrument demonstrates appropriate targeting and measurement precision (Tennant & Conaghan, 2007). Furthermore, Rasch analysis provides detailed information regarding item level performance and scale functioning that cannot be obtained through traditional reliability analyses alone (Petrillo et al., 2024).

Accordingly, the theoretical framework of the present study posits that healthy dietary behavior is a latent construct that can be measured through the REAP-S and that an Arabic adaptation should demonstrate conceptual equivalence, structural validity, internal consistency, and satisfactory measurement properties. The integration of cross-cultural adaptation theory, Classical Test Theory, and Rasch Measurement Theory provides the conceptual foundation for evaluating whether the Arabic REAP-S is a valid and reliable instrument for assessing dietary behaviors among Arabic-speaking young adults.

## Method

### 2.1 Study Design and Participants

The Rapid Eating Assessment for Participants–Shortened Version is a previously published dietary screening instrument developed to assess eating behaviors and dietary practices. The English-language REAP-S has previously undergone evaluation of its reliability and validity (Shankar et al., 2023). The instrument was not developed by the authors of the present study. Rather, the present study aimed to translate, culturally adapt, and evaluate the psychometric properties of the REAP-S in Arabic among Arabic-speaking young adults.

It evaluated construct validity, internal consistency, item level functioning, and overall measurement properties using Confirmatory Factor Analysis (CFA) and Rasch Measurement Theory. The study population comprised undergraduate and graduate nursing students from several universities in Saudi Arabia during the 2025–2026 academic year. The participating universities were involved in participant recruitment, with close cooperation from the administrative offices. After obtaining institutional approval, the researchers checked students’ enrollment records to determine whether any nursing students were enrolled at different academic program levels.

The eligible participants had to be full time nursing students who could understand either Arabic or English at the time of enrollment. Official university communications (email and Blackboard announcements) were used to send invitations to participate. An electronic consent form, a distinct survey URL, and an information page regarding the study were sent with the invitations. To boost involvement, school personnel used school communication channels and made in class announcements to aid recruitment. Digital posters and study invitations were distributed on student portals and other media platforms to bolster this. An online survey that was disseminated via official university communication channels was used to gather data between September 2025 and February 2026. After giving their electronic informed consent, eligible students completed the survey.

### 2.2 The sampling and sample size

Those who volunteered to take part in the study throughout the recruiting period were recruited using convenience sampling. The conventional formula for predicting proportions, a conservative estimate (p = 0.50), a 98% confidence level, and a 5% margin of error were used to get the necessary sample size. An extra 15% was added to account for possible nonresponse or incomplete data, resulting in a total sample size of roughly 800 individuals.

The number of participants in relation to the number of items on the 52-item factor-analyzed instrument used in this study was utilized to establish sample adequacy. The final sample comprised 500 nursing students with 9.6 individuals per item, which was suitable for the Rasch analysis and confirmatory factor analysis that were intended.

### 2.3 Translation, Content, and Face Validity, and Piloting

To ensure language and conceptual equivalency, the translation of REAP-S was based on a standardized forward backward translation technique. Two bilingual translators independently translated this from the original English into Arabic. The instrument was then translated into Arabic and back into English in a reconciled version by two separate translators who were blind to the original instrument. To make sure the translations were conceptually and semantically equivalent, the study team compared the original and back-translations.

A panel of professionals in public health, nursing, and psychometrics verified the content validity. The items’ cultural appropriateness, relevance, and clarity were assessed by the experts. Minor modifications were made to the items without altering their original meaning to make them as written as feasible and to make sure they are appropriate for the situation.

A small sample of nursing students who were representative of the target demographic evaluated the instrument’s readability, comprehensibility, and clarity to test its face validity. They suggested a few small adjustments, which were implemented. After that, a tiny sample of this Arabic document was pilot tested to assess its viability, readability, and completion time. According to the initial findings, the instrument was simple to understand and didn’t require many modifications.

### 2.4 Tool Validation

Internal consistency of the 13-item core diet questionnaire was evaluated using Cronbach’s alpha. Construct validity was examined by testing whether the originally proposed single factor structure of the diet questionnaire provided an adequate fit to the current data. Confirmatory factor analysis was used to test the proposed model structure, whereas Rasch analysis was conducted to evaluate the instrument’s ability to differentiate levels of dietary behaviour and to assess item hierarchy.

Confirmatory factor analysis was performed to test the fit of the proposed single-factor model for the 13-item core diet questionnaire. Model fit was evaluated using the Comparative Fit Index (CFI), Tucker-Lewis Index (TLI), root mean square error of approximation (RMSEA), and standardized root mean square residual (SRMR). Internal consistency for the overall scale was examined using Cronbach’s alpha.

Unidimensional Rasch analysis for polytomous responses was conducted using the partial credit model (Bond & Fox, 2015). Person reliability was examined using expected a posteriori and weighted likelihood estimates. Infit and outfit statistics were obtained, with mean square values between 0.50 and 1.50 considered acceptable. Item thresholds were inspected, and a Wright map was generated to evaluate person-item targeting.

### 2.5 Statistical Analysis

The data were analyzed in R version 4.5.3(Team, 2020), with the packages psych(Psych Revelle, 2023), lavaan(Rosseel, 2012), and TAM(Robitzsch et al., 2020). Categorical variables were summarized as frequencies and percentages, while continuous variables were presented as medians (IQR). A p-value of < 0.05 was considered statistically significant.

#### Confirmatory Factor Analysis

The model from the original factor of REAP-S I was tested on the current data through confirmatory factor analysis to check the model fit. The Comparative Fit Index (CFI), Tucker-Lewis Index (TLI), root mean square error of approximation (RMSEA), and standardized root mean square residual (SRMR) were used to test model fit. Cronbach’s alpha was used to assess internal consistency for each factor.

#### Rasch Analysis

The polytomous responses were analyzed with multidimensional Rasch analysis, using the partial credit model(Bond & Fox, 2013). A unidimensional model and a six-dimensional model parallel to the original REAP-S domains were estimated, and the latter was retained because it yielded better information criteria. We used expected a posteriori (EAP) and weighted likelihood estimates (WLE) to assess person reliability. Infit and outfit statistics were calculated, and mean square values ranging from 0.50 to 1.50 were deemed acceptable. An inspection of item thresholds and a Wright map to assess person-item targeting was conducted. Differential item functioning (DIF) by sex was also analyzed as a sensitivity analysis with the construction of the item-by-sex interaction terms. The absolute values larger than 1.96 were considered as raw evidence of DIF, and Holm adjustment was performed to control for multiple testing.

### 2.6 Ethical Considerations

The study was conducted in accordance with the ethical principles of the Declaration of Helsinki. Participation was entirely voluntary, and written informed consent was obtained from all participants before data collection. Participants were informed about the purpose of the study, their right to refuse participation or withdraw at any time without any consequences, and the confidentiality and anonymity of their responses. No identifiable personal information was collected, and all data were securely stored and accessible only to the research team. During a designated data collection time, the online questionnaire was posted using Google Forms. Interested students accessed the survey using the secure electronic link provided in the study invitation.

The students had already read the consent form electronically, which described the purpose, voluntary nature, and confidentiality of data for this study. By clicking the “I agree” box at the start of the survey, the participants expressed their approval. Those who selected “I don’t agree” were processed further, but no information was collected from them. Every piece of information collected was kept confidential and anonymous. No risks were anticipated for study participants, and their reputation or position would not be compromised if they declined to take part.

## Results

The sociodemographic characteristics of the participants included in the psychometric analyses are displayed in Table 1. The final analytic sample comprised 490 participants, of whom 265 (54.1) were female. The median age was 20 years (interquartile range 19-22).

**Table 1.** Sociodemographic characteristics of the study participants.

| Characteristic | Value |
| --- | --- |
| Age, years | 20 (19-22) |
| Sex |  |
| Female | 265 (54.1) |
| Male | 225 (45.9) |
| Third year | 49 (10.0) |
| University type |  |
| Private | 23 ( 4.7) |
| Public | 467 (95.3) |
| Economic status |  |
| Low | 42 ( 8.6) |
| Moderate | 415 (84.7) |
| High | 33 ( 6.7) |
| Smoking status |  |
| Current smoker | 75 (15.3) |
| Former smoker | 27 ( 5.5) |
| Never smoked | 388 (79.2) |
| Chronic disease or disability |  |
| No | 472 (96.3) |
| Yes | 18 ( 3.7) |

*Note. Continuous data are presented as median (25th-75th percentiles), and categorical data are* *presented as count (%).*

### Tool Validation

To assess the validity of the proposed single-factor model, CFA was conducted. The initial 13-item, one-factor model showed strong model fit (CFI = 0.984, TLI = 0.978, RMSEA = 0.076, and SRMR = 0.061). The highest standardized factor loading was observed for Item 5. Eat <2 servings vegetables daily (0.768), while the lowest was observed for Item 1. Skip breakfast (0.429). Cronbach’s alpha for the 13-item scale was acceptable at 0.863. The standardized factor loadings are presented in Table 2.

**Table 2.** Diet questionnaire item factor loadings, outfit and infit mean square statistics, and threshold parameters.

| Item | Loading<br>(SE) | Outfit<br>MSQ | Infit<br>MSQ | Threshold<br>1 | Threshold<br>2 | Median<br>score<br>(IQR) |
| --- | --- | --- | --- | --- | --- | --- |
| Item 1. Skip breakfast | 0.429<br>(0.043) | 1.311 | 1.283 | -1.747 | 0.765 | 2 (2.00-3.00) |
| Item 2. Eat 4+ fast-food meals per week | 0.476<br>(0.040) | 1.209 | 1.194 | -0.635 | 1.897 | 2 (1.00-2.00) |
| Item 3. Eat <2 servings whole grains daily | 0.705<br>(0.029) | 0.934 | 0.944 | -1.469 | 1.574 | 2 (2.00-2.00) |
| Item 4. Eat <2 servings fruit daily | 0.734<br>(0.025) | 0.869 | 0.903 | -1.508 | 1.220 | 2 (2.00-3.00) |
| Item 5. Eat <2 servings vegetables daily | 0.768<br>(0.023) | 0.826 | 0.866 | -1.358 | 1.219 | 2 (2.00-3.00) |
| Item 6. Consume <2 servings milk/yogurt/cheese daily | 0.750<br>(0.024) | 0.913 | 0.944 | -1.150 | 1.333 | 2 (1.00-2.00) |
| Item 7. Eat >8 ounces of meat/chicken/fish daily | 0.581<br>(0.035) | 1.107 | 1.099 | -1.373 | 1.220 | 2 (2.00-3.00) |
| Item 8. Use processed instead of fresh meats | 0.718<br>(0.027) | 0.883 | 0.907 | -0.967 | 1.852 | 2 (1.00-2.00) |
| Item 9. Eat fried foods | 0.602<br>(0.034) | 1.027 | 1.038 | -1.625 | 1.126 | 2 (2.00-3.00) |
| Item 10. Eat salty snacks/chips regularly | 0.629<br>(0.033) | 0.938 | 0.976 | -1.019 | 1.433 | 2 (1.00-2.00) |
| Item 11. Add fats to bread/rice/potatoes at table | 0.649<br>(0.031) | 0.962 | 0.973 | -1.217 | 1.471 | 2 (1.00-2.00) |
| Item 12. Eat sweets >2 times daily | 0.605<br>(0.032) | 1.041 | 1.041 | -1.026 | 1.509 | 2 (1.00-2.00) |
| Item 13. Drink >16 oz sugar-sweetened beverages daily | 0.612<br>(0.032) | 0.941 | 0.978 | -0.914 | 1.791 | 2 (1.00-2.00) |

*Note. Item rows display standardized factor loadings from the final confirmatory factor analysis, item-level outfit and infit mean square values from the unidimensional partial credit model, the two operational threshold parameters, and the observed median item score with interquartile range.*

### Rasch Model

The unidimensional Rasch model was suitable for the 13-item diet scale. The reliability values based on expected a posteriori (EAP) and weighted likelihood estimates (WLE) were 0.872 and 0.848, respectively. Table 2 displays the infit and outfit values, supporting item fit within the Arabic diet questionnaire and the instrument’s ability to differentiate between varying levels of dietary behavior among participants.

The infit and outfit mean square values were within the acceptable range for all 13 items. The item thresholds demonstrated variation in item difficulty, with correctly ordered thresholds across all items. The lowest threshold was the first threshold of Item 1. Skip breakfast (-1.747), while the highest threshold was the second threshold of Item 2. Eat 4+ fast-food meals per week (1.897). The highest outfit and infit values were observed for Item 1. Skip breakfast (1.311 and 1.283, respectively), although these remained within the acceptable range. The median total score of the 13-item scale was 26 (23.00-28.00) out of a maximum possible score of 39. Table 2 presents the item thresholds and observed item medians. The Wright map indicated acceptable person-item targeting (Figure 1).

**Figure 1.**
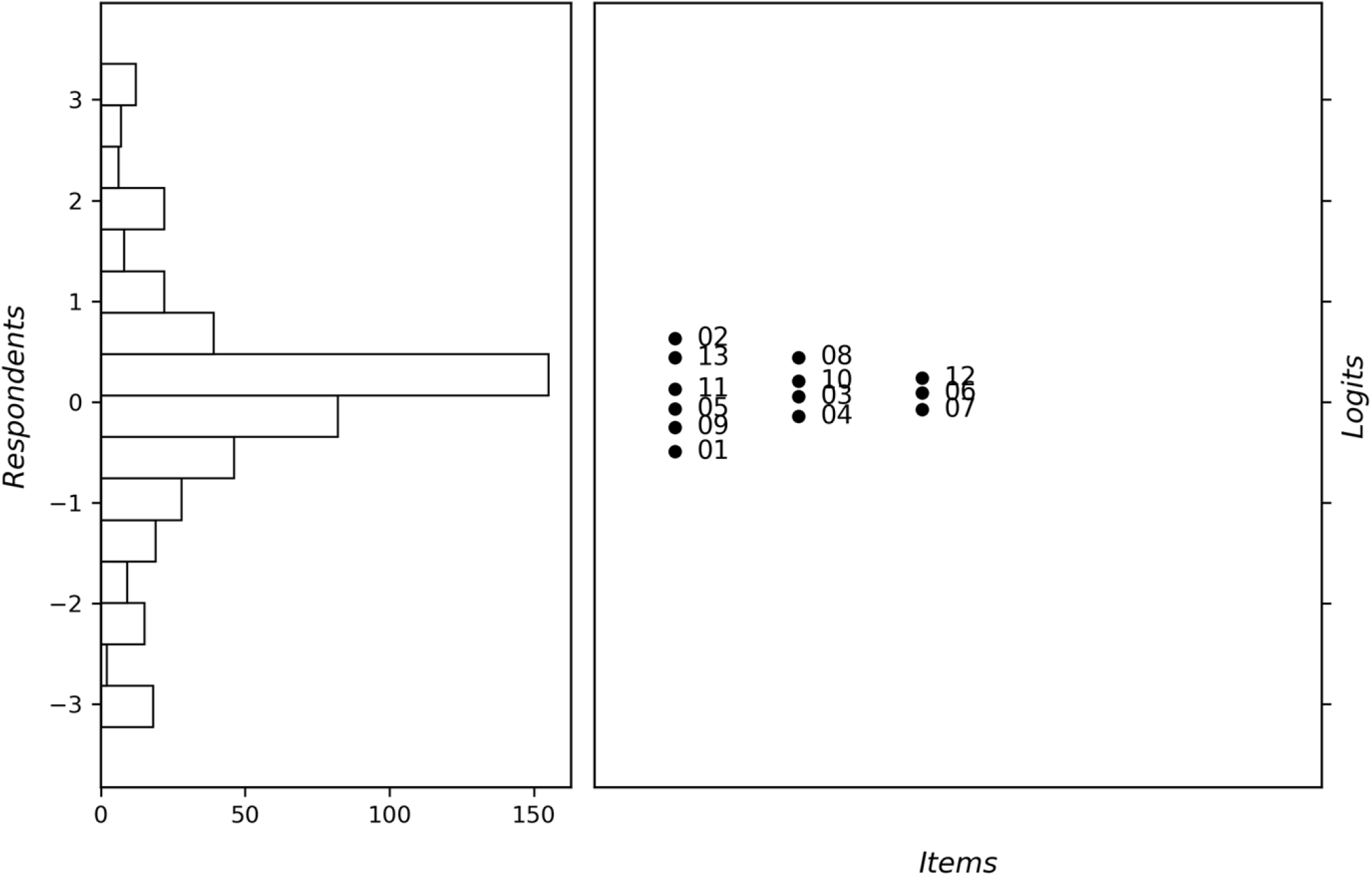
Wright map of the Arabic 13-item core diet questionnaire. *Note. The left panel displays the person distribution, and the right panel displays the item locations on the same latent trait scale. Item numbers correspond to the original item sequence in the 13-item core questionnaire*.

## Discussion

The present study provides robust evidence supporting the validity and reliability of the Arabic version of the Rapid Eating Assessment for Participants–Shortened Version (REAP-S). The use of both Confirmatory Factor Analysis (CFA) and Rasch Measurement Theory strengthens the psychometric evaluation by combining traditional and modern measurement approaches, which is increasingly recommended in scale validation research(Boateng et al., 2018; Tennant & Conaghan, 2007).

The CFA results confirmed the unidimensional structure of the Arabic REAP-S, with excellent fit indices (CFI = 0.984, TLI = 0.978, SRMR = 0.061). These findings are consistent with the original conceptualization of REAP-S as a single-factor dietary screening tool designed to capture overall dietary quality rather than isolated food components (Segal-Isaacson et al., 2004). Similar psychometric performance has been reported in updated versions of the tool, further supporting its structural stability across populations(Shankar et al., 2023).

Although the RMSEA value (0.076) was slightly above the strict threshold, it still indicates acceptable model fit, particularly in models with a relatively small number of items and large sample sizes, where RMSEA can be overly sensitive(Kenny et al., 2015). Overall, the CFA results support that the Arabic REAP-S adequately represents a single latent construct of dietary behavior.

Factor loadings showed that vegetable intake had the strongest association with the latent construct, suggesting that it is a central indicator of dietary quality in this population. This aligns with global dietary guidelines emphasizing vegetables as a core component of healthy eating patterns(WHO, 2020). In contrast, breakfast skipping showed the weakest loading, which may reflect changing dietary behaviors among young adults, including intermittent fasting practices and irregular eating schedules that are not necessarily indicative of poor overall diet quality.

Internal consistency was good (Cronbach’s alpha = 0.863), exceeding the commonly accepted threshold of 0.70 for research instruments (Nunnally & Bernstein, 1994). This indicates that the Arabic REAP-S items are sufficiently correlated while still capturing multiple dimensions of dietary behavior without redundancy.

Rasch analysis further supported the psychometric strength of the instrument. Both EAP (0.872) and WLE (0.848) reliability coefficients indicated good measurement precision across different levels of dietary behavior. This supports the instrument’s ability to differentiate between individuals with healthier and less healthy dietary patterns, which is essential for screening tools used in population health settings(Bond & Fox, 2013).

All items demonstrated acceptable infit and outfit statistics, indicating good model-data fit and supporting the unidimensional structure identified in CFA. This consistency between CFA and Rasch results provides strong convergent evidence for construct validity, which is considered best practice in modern psychometric validation studies(Boateng et al., 2018).

The ordered thresholds across all items suggest that participants were able to distinguish clearly between response categories. This is an important finding in cross-cultural adaptation studies, as poorly translated response scales often result in threshold disordering due to semantic or cultural misunderstandings(Lin et al., 2025). The absence of such issues in the present study indicates successful linguistic and cultural adaptation of the instrument.

Item difficulty analysis revealed meaningful variation across dietary behaviors. Skipping breakfast was the easiest behavior to endorse, suggesting it is highly prevalent among participants, whereas frequent fast-food consumption represented a more severe dietary behavior. This pattern is consistent with dietary trends observed in university populations, where meal skipping is common but frequent fast-food intake varies more widely(Deliens et al., 2014; Pendergast et al., 2016).

The Wright map demonstrated acceptable person-item targeting, indicating that the difficulty of the items appropriately matched the dietary behavior levels of the sample. Good targeting improves measurement precision and reduces bias in score interpretation, supporting the suitability of the Arabic REAP-S for use in young adult populations(Bond & Fox, 2013).

From a public health perspective, the availability of a validated Arabic dietary screening tool is highly significant. Non-communicable diseases such as obesity, diabetes, and cardiovascular disease continue to rise in the Middle East and North Africa region, where dietary behaviors are a major contributing factor(Collaborators, 2020). Brief and reliable tools such as REAP-S can facilitate early identification of poor dietary habits in clinical and community settings. The integration of CFA and Rasch methodologies represent a major strength of this study.

## Conclusion

The Arabic version of the Rapid Eating Assessment for Participants–Shortened Version (REAP-S) demonstrated strong psychometric properties in the present study, supporting its use as a valid and reliable instrument for assessing dietary behaviors among Arabic speaking adults. The findings from both Confirmatory Factor Analysis and Rasch Measurement Theory consistently supported a unidimensional structure, with acceptable model fit, strong internal consistency, and well functioning item characteristics, in line with contemporary recommendations for the validation of patient-reported outcome measures and health assessment instruments (Mokkink et al., 2023; Furr, 2021). The absence of significant item misfit and the presence of ordered response thresholds further confirmed the adequacy of the cultural and linguistic adaptation process, supporting current guidelines for cross-cultural adaptation and validation studies (Boateng et al., 2024).

In addition, the instrument showed good measurement precision and appropriate person–item targeting, indicating its suitability for screening dietary behaviors in young adult populations. These findings are consistent with recent applications of Rasch Measurement Theory, which emphasize the importance of targeting, reliability, and response category functioning in evaluating measurement quality (Petrillo et al., 2024; Boone et al., 2021). The convergence of results from both classical and modern psychometric approaches strengthens confidence in the robustness of the Arabic REAP-S and supports its use in both research and applied settings (Mokkink et al., 2023).

Given the growing burden of diet related non-communicable diseases in Arabic speaking countries, the availability of a brief, culturally adapted, and psychometrically sound dietary assessment tool is of significant public health value (World Health Organization, 2024). The Arabic REAP-S can be effectively used in clinical practice, community health programs, and epidemiological research to rapidly identify individuals with suboptimal dietary patterns and guide targeted nutritional interventions, consistent with current recommendations for dietary assessment and preventive healthcare strategies (Afshin et al., 2023).

Future research should further examine the tool’s applicability across different age groups, clinical populations, and cultural contexts within the Arab region to establish broader generalizability and measurement invariance (Mokkink et al., 2023). Longitudinal studies are also warranted to assess the instrument’s responsiveness to dietary changes over time and its utility in evaluating the effectiveness of nutrition interventions, as recommended in contemporary psychometric research (Furr, 2021; Petrillo et al., 2024).

## Statement and Declaration

### Ethical approval and consent to participate

This study was conducted in accordance with the Declaration of Helsinki, and ethical approval for this study was obtained from the Research Ethics Committee (REC) at the University of Hail, Saudi Arabia (Ethical Approval No: H-2025-954). Written informed consent was obtained from all participating individuals before their enrollment in the study.

### Consent for Publication

Not applicable.

### Data Availability statement

The datasets generated and/or analyzed during the current study are not publicly available due to institutional privacy policies and ethics considerations regarding participant confidentiality but are available from the corresponding author on reasonable request, subject to a data sharing agreement.

### Research funding

This research received no specific grant from any funding agency in the public or commercial.

### No Patient or Public Contribution

Patients and members of the public were not involved in the design, conduct, reporting, or dissemination of this methodological validation study. Participants contributed only through completion of the study questionnaire.

### Authors’ contributions

T M.A-B, W.A-Q, S N.A-Q, A Y.A-H, and O.A-K: Conceptualization, methodology, formal analysis, validation, investigation, data curation, writing of the original draft, visualization, and subsequent writing, reviewing, and editing. All authors read and approved the final manuscript for publication.

## Acknowledgment

During the preparation of this work, the author(s) used ChatGPT by OpenAI to improve the readability and language of the manuscript. After using this tool, the author(s) carefully reviewed and edited the content as needed and take full responsibility for the content of the published article.

